# Sleepless Nights: A Single-Center Cross-Sectional Study Reveals Gender-Related Sleep Quality in Dialysis Patients

**DOI:** 10.1101/2025.09.24.25336589

**Authors:** Yan Liu, Yumei Liao, Yanmei Peng, Yuhan Diao, Guang Yang

**Author notes:** Correspondence: Guang Yang. Contributed equally to this work (Yan Liu, Yumei Liao). Other emails: Yan Liu, Yumei Liao, Yanmei Peng, Yuhan Diao.

## Abstract

**Background:** Poor sleep quality (PSQ) is prevalent among patients on dialysis. However, the etiology and coping strategies for PSQ may vary based on gender. The influence of gender differences remains unexplored. This study investigates the influence of gender on PSQ and associated risk factors in patients on dialysis.

**Methods:** This study included 260 patients who underwent dialysis at Peking University Shenzhen Hospital, China between May 2023 and October 2023. Patient health, sleep patterns, and emotional status were assessed using structured questionnaires. Blood composition is determined via the most recent medical examination record. Both univariate and multivariate analyses were employed to identify factors influencing sleep quality across genders. Additionally, the Student’s t-test was used to identify factors that cause differences in sleep quality between genders.

**Results:** In general, compared with male patients, female patients presented inferior sleep quality. Comprehensive factor analysis revealed that male PSQ was predominantly associated with pruritus, anxiety, and decreased ferritin levels. Conversely, female PSQ was associated with restless legs syndrome, elevated systolic blood pressure, and depression.

**Conclusions:** Gender-specific risk factors are evident for PSQ among patients on dialysis, emphasizing the need for personalized treatment and tailored nutritional interventions. This study has significant implications for nephrology nursing.

## 1 Introduction

Poor sleep quality (PSQ) is a subjective perception of sleep that is unsatisfactory or fails to meet an individual’s expectations and can lead to feelings of fatigue, irritability, and impaired daytime functioning [1, 2]. It is particularly prevalent in patients with chronic kidney disease (CKD) [3, 4], with a reported incidence of 78.2% in those with more severe disease [5–7]. CKD refers to a long- term condition characterized by the gradual loss of kidney function over time[8]. The progressive nature of CKD can lead to a decline in the kidney’s ability to filter waste products and excess fluids from the body, resulting in the accumulation of toxins and fluid imbalances [9]. Sometimes, CKD patients are also called patients on dialysis because they need dialysis for a long time to stay alive. Owing to the complexity of its etiology, effectively improving CKD-related PSQ has been a clinical challenge.

The sleep quality of inpatients is a key concern for both medical staff and nurses. In the aspect of nursing, nurses are chiefly responsible for basic nursing tasks. They create a comfortable ward environment and assist patients in maintaining a comfortable posture. Meanwhile, they closely monitor patients’ conditions and promptly address symptoms that impact sleep. Besides, nurses offer psychological support to alleviate patients’ negative emotions and carry out sleep-related health education to help patients form good sleep habits. Doctors, in contrast, are tasked with treating the root causes of diseases to fundamentally resolve sleep disorders triggered by illnesses. They rationally administer drug therapies to enhance patients’ sleep. Moreover, doctors conduct a comprehensive evaluation of patients’ overall situations, assess the influence of sleep problems on patients’ conditions, and promptly adjust treatment strategies. Given that nurses have more frequent contact with patients, if they detect that a patient has low-quality sleep, they should report it to the doctor without delay. For nursing in the nephrology department, timely nursing interventions play a pivotal role in improving the sleep quality of nephrology patients.

The etiology of CKD-related PSQ is complex, involving physiological, psychological, and lifestyle factors, making treatment challenging. While PSQ has been widely studied in the general population, less is known about its specific impact on CKD patients. Addressing sleep disturbances in this group is crucial, as poor sleep is linked to worsened health outcomes like cardiovascular disease, increased mortality, and mental health decline. This underscores the need for further investigation, particularly into gender differences in CKD-related PSQ. Research indicates that females may be more susceptible to insomnia, excessive dreaming, and sleepiness than males are due to hormonal changes related to their biological cycle, pregnancy, and menopause [10, 11]. On the other hand, males have a greater risk of developing sleep apnea syndrome, which may be linked to differences in fat and muscle distribution around the neck [12]. Coping mechanisms also vary between genders, with females being more likely to seek support than males [13].

It is unclear whether there are also gender differences in CKD-related PSQ. We hypothesized that gender differences in CKD-related PSQ exist, requiring a more tailored approach to healthcare for male and female patients. Therefore, this study aimed to explore these differences through a single-center, cross-sectional study, analyzing data from 260 patients undergoing dialysis (151 males and 109 females). Utilizing both univariate and multivariate analysis, we sought to identify gender- specific risk factors and propose a more individualized model for managing CKD-related PSQ.

## 2 Methods

### 2.1 Ethics and procedures

The research protocols were scrutinized and received approval from the Ethics Committee of Peking University Shenzhen Hospital (Approval No. 2023--066). Before initiation, all participants were verbally informed about the study’s objectives and significance, and their informed consent was duly procured. All patients signed a pan-informed consent form. The Strengthening the Reporting of Observational Studies in Epidemiology (STROBE) statement was followed in the planning, implementation, and reporting of the present work [14]. The workflow is described in Figure 1. This was a single-center cross-sectional study. Data collection included questionnaire surveys and a retrospective analysis of clinical data. The study ensured that there was no harm to the participating patients.

**Figure 1.**
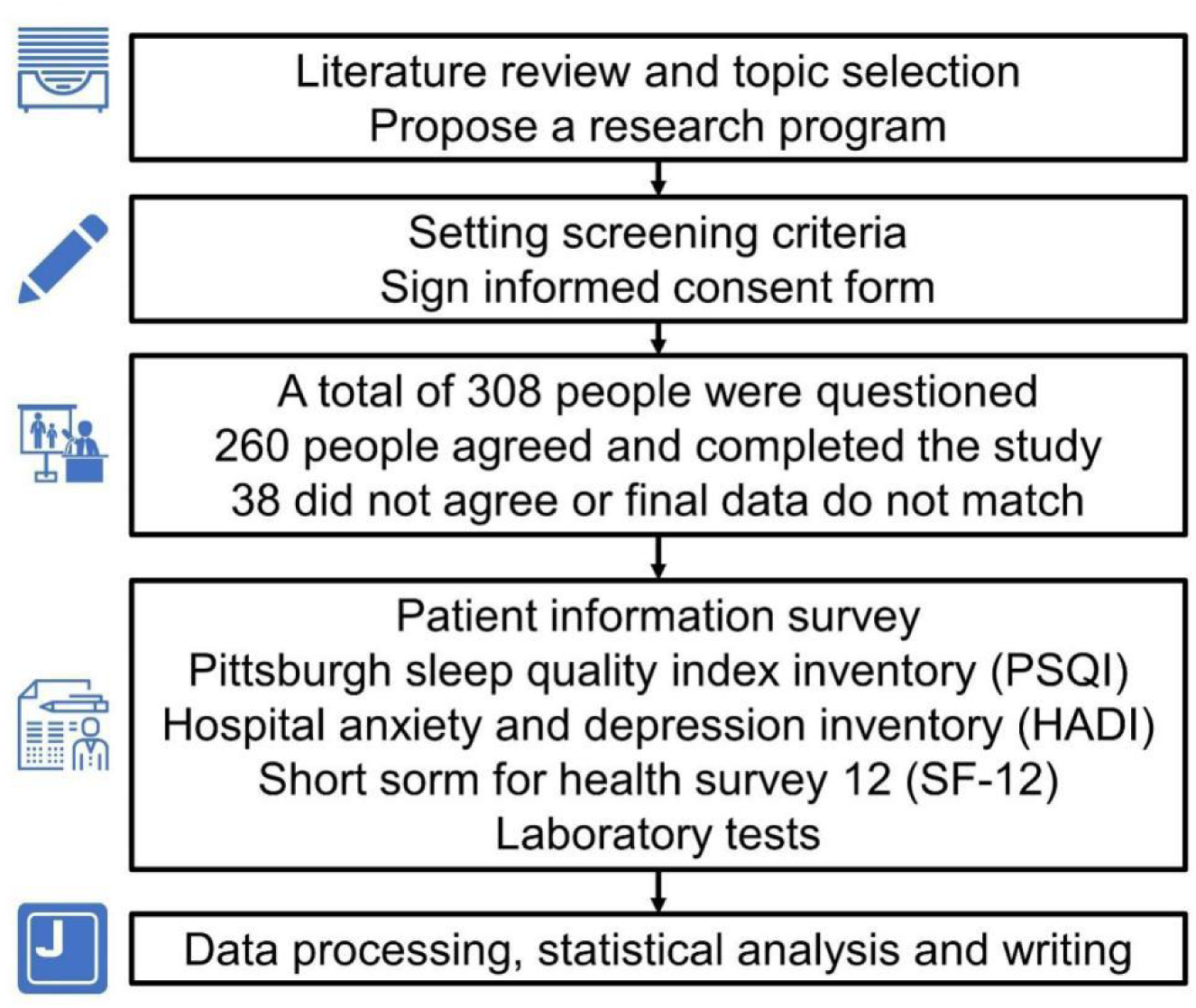
Research flowchart.

### 2.2 Patient information

The target population for the survey consisted of patients who were receiving dialysis at Peking University Shenzhen Hospital between May 2023 and October 2023. Typically, these patients have severe CKD and rely on dialysis treatment 2-3 times weekly, which results in prolonged contact with healthcare professionals and increased cooperativeness. The inclusion criteria were as follows: (i) regular dialysis treatment duration >3 months; (ii) age range of 18-90 years; (iii) literacy and effective communication capabilities with health care personnel; and (iv) comprehension of the context of this study, voluntary participation, and provision of informed consent. The exclusion criteria were as follows: (i) prevalent communication impediments and (ii) recent acute complications necessitating unhampered recovery. A total of 260 study subjects were included, with 130 patients each on peritoneal dialysis (PD) and hemodialysis (HD) to neutralize the effect of the HD modality. See the supplementary materials for more information.

### 2.3 Blood tests

For CKD patients in a stable phase, quarterly blood tests are recommended. Typically, blood samples are drawn from an elbow vein and stored either in anticoagulation tubes (#101680720, EDTA-K2-2.0 mg/mL, Ivory, China) for standard blood panels or in procoagulation tubes (Separation Gel Coagulation Tube 3.5 mL, Ivory, China) for other blood samples. The testing center utilized standard assays: routine blood analyses were performed using the Macon SYSMEX, followed by blood biochemical indicators analysis using Beckmann AU5800, and parathyroid function test using Beckmann DXI800. The results of the most recent test were used as patient blood information in this investigation.

### 2.4 Health status

Patient health was evaluated using the Health Survey 12-item Short Form (SF-12) [15], which was completed by all participants independently and with a uniform prompt to ensure full comprehension. The questionnaire typically took approximately 15 minutes to complete and was collected by the researcher upon completion. The participants were reminded to make corrections if any obvious errors were found.

### 2.5 Sleep quality

Patient sleep was evaluated through the Pittsburgh Sleep Quality Index (PSQI) [16], which encompasses seven factors: subjective sleep quality (SSQ), sleep latency (SL), sleep duration (Sdu), habitual sleep efficiency (HSE), sleep disturbance (Sdi), daytime dysfunction (DD), and the use of sleep medication (USM). Each factor was rated on a 0-3 scale, and the sum of all factor scores constituted the total PSQI score. Higher scores on the PSQI indicate poorer sleep quality. A total PSQI score greater than 5 was used as the critical threshold to identify low sleep quality.

### 2.6 Lifestyle

The patients’ alcohol, tea, and coffee consumption was recorded, and their intake was assessed primarily on a weekly basis. A score of -3 was assigned for no intake, 1 for once a week, 2 for 2--3 times a week, and 3 for 4--7 times a week. Additionally, if the intake of alcohol, tea, or coffee exceeded 50 ml, 500 ml, or 200 ml per day, respectively, they were considered heavy drinkers and were rated as 4. Exercise was also documented, primarily consisting of jogging, brisk running, walking, cycling, and swimming. If the patient did not exercise in the previous year, it was marked as -3. Inactivity was scored as 0, once a week as 1, 2--3 times per week as 2, and 4--7 times per week as 3. Walking for more than 40 minutes was considered one valid exercise session, and other single sessions needed to be longer than 20 minutes.

### 2.7 Emotional state

The Hospital Anxiety and Depression Inventory (HADI) was used to evaluate the anxiety and depression status of the patients [17]. It consists of seven items, each for anxiety and depression. The score interpretations were as follows: 0-7 (asymptomatic), 8-10 (borderline), and 11-21 (explicitly symptomatic).

### 2.8 Statistics

The results are presented as the means ± standard deviations (SDs), and the count data are presented as frequency‒composition ratios. R Studio 4.2.2 was used for the statistical analyses, with simple linear regression applied to identify factors influencing sleep quality by sex. Factors scoring *p* ≤ 0.05 by univariate analysis were further analyzed via multivariate analysis and variable screening via stepwise regression. The final multiple linear regression model was used to determine the factors influencing patients’ sleep quality. The unpaired Student’s t-test facilitated statistical comparisons at the sex level, with *p* ≤ 0.05 indicating statistical significance. *, *p* ≤ 0.05. **, *p* ≤ 0.01.

## 3 Results

### 3.1 Female patients experience poorer sleep quality than males do

To evaluate the influence of personal demographics on sleep quality among patients of different genders, we analyzed age, education, marital status, work status, and economic status among 260 patients on dialysis (Table S1). Among these participants, 151 (58.1%) were male, and 109 (41.9%) were female. The average age of the male patients was 52.21 ± 13.03 years, whereas the average age of the female patients was 50.50 ± 13.29 years. A two-sample t-test indicated that the mean PSQI score for male patients (6.95 ± 3.63) was significantly lower than that for female patients (8.44 ± 4.16), with a statistically significant difference [95% CI: (-2.471, -0.515), *p* ≤ 0.0029]. These findings indicate that females generally experience poorer sleep quality than males do.

### 3.2 Univariate analysis of factors affecting sleep quality

#### 3.2.1 PSQ is associated with medical conditions

We investigated the relationships between sleep quality and various medical parameters, including primary disease, type of dialysis, skin itching episodes, blood pressure, and pulse rate (Table 1). Male patients with primary renal disease and recent frequent episodes of skin itching were found to have compromised sleep quality. Among female patients, those undergoing hemodialysis experienced more sleep disturbances than did those on peritoneal dialysis. Additionally, restless leg symptoms (RLS) are more prevalent as a cause of sleep disturbance in women. A notable positive correlation emerged between systolic blood pressure and overall PSQI score among females, indicating that they are more likely to experience poorer sleep quality with more sleep disturbances. These findings also suggest that maintaining optimal blood pressure control should be an important consideration for improving PSQ in female patients.

**Table 1.**
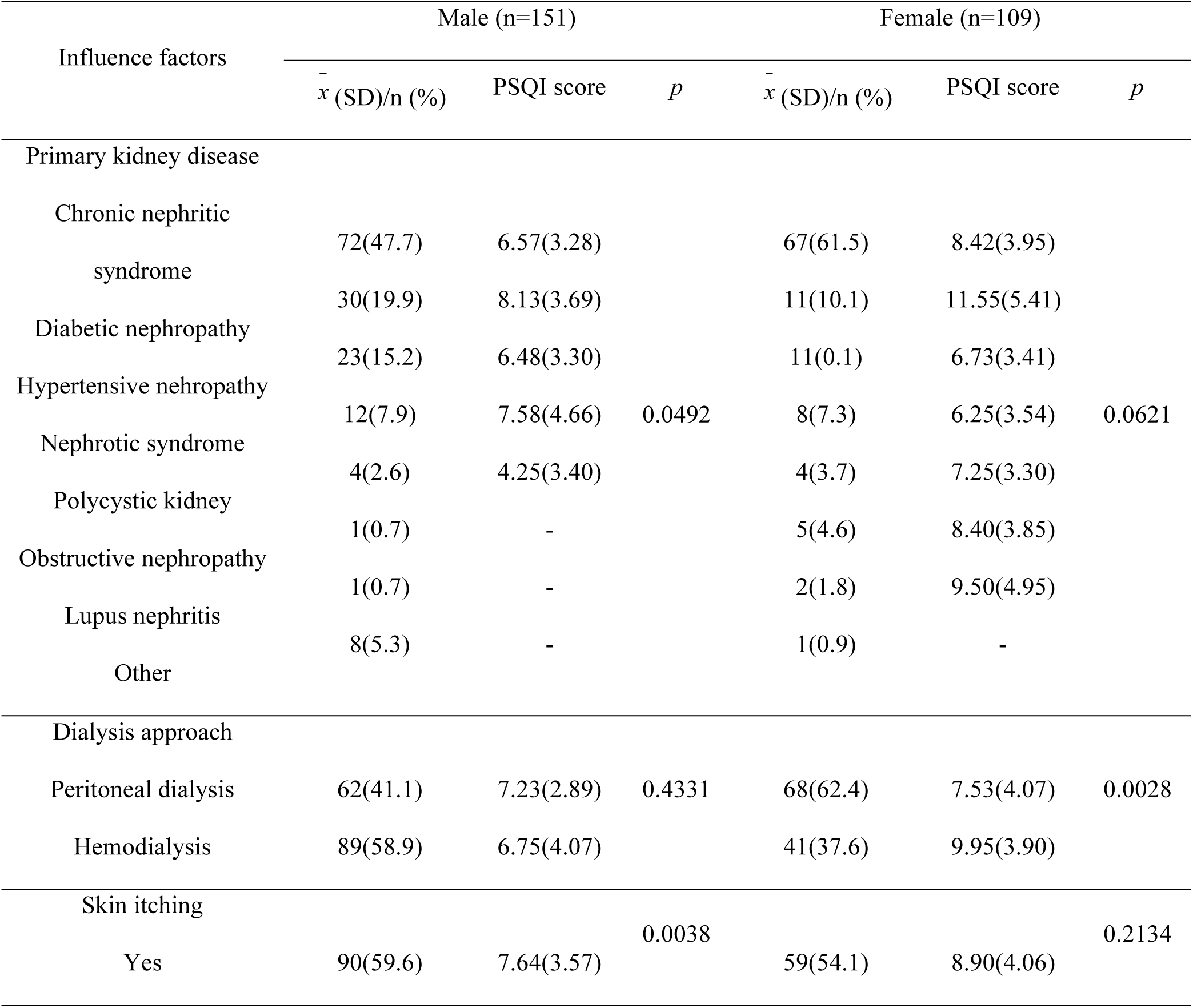
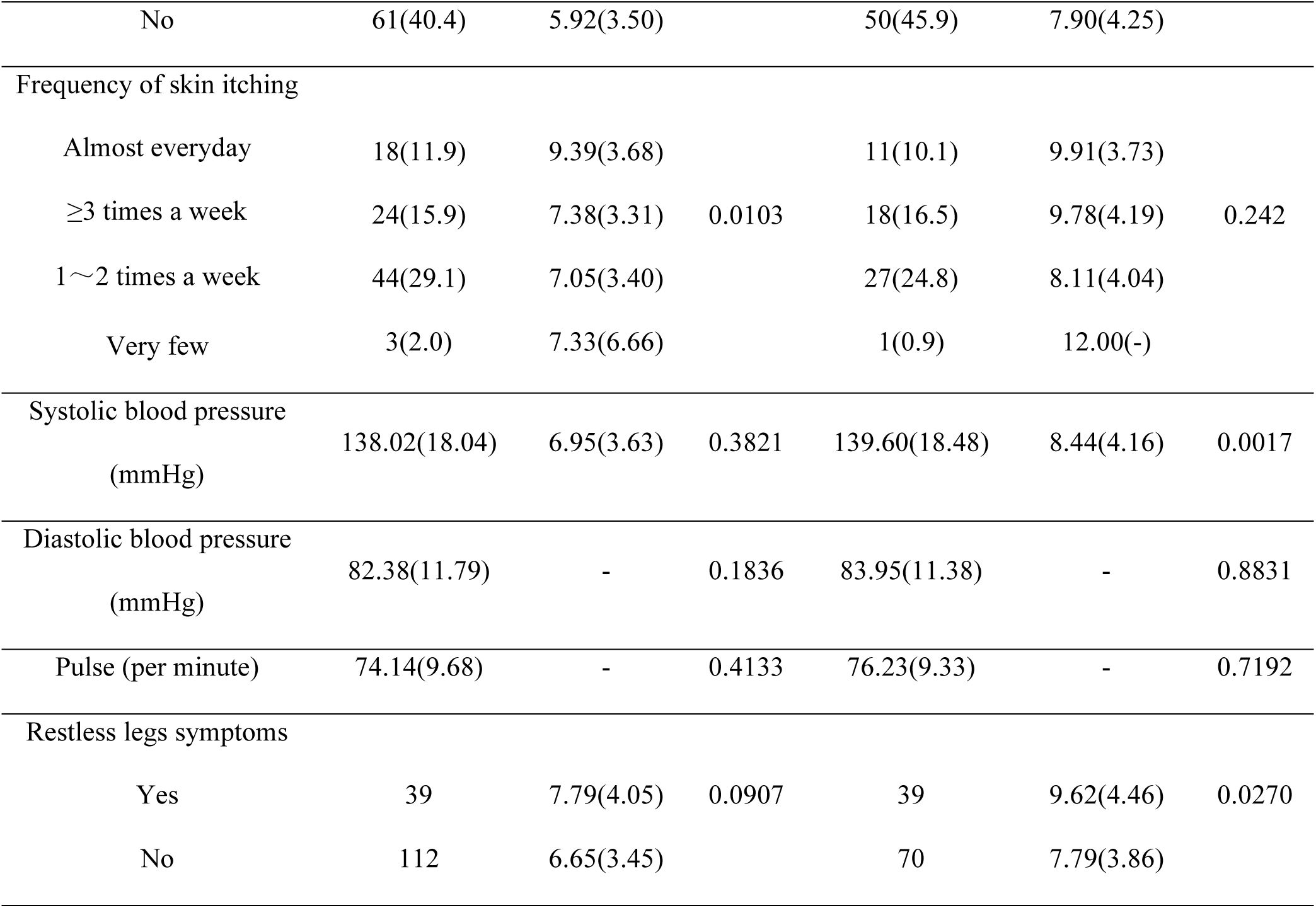
Univariate analysis of disease and sleep quality.

#### 3.2.2 PSQ is associated with emotional well-being

To discern the relationship between emotional states and sleep quality, we examined the levels of anxiety and depression among the patients (Table 2). Upon categorizing individuals on the basis of their rating scale scores, we found evidence demonstrating a significant positive association between anxiety and depressive symptoms and sleep disturbances. Specifically, individuals with more severe anxiety and depression presented higher total PSQI scores. These findings suggest that greater psychological distress is associated with severe PSQ in patients on dialysis. Treating underlying mood disorders could help improve sleep quality.

**Table 2.**
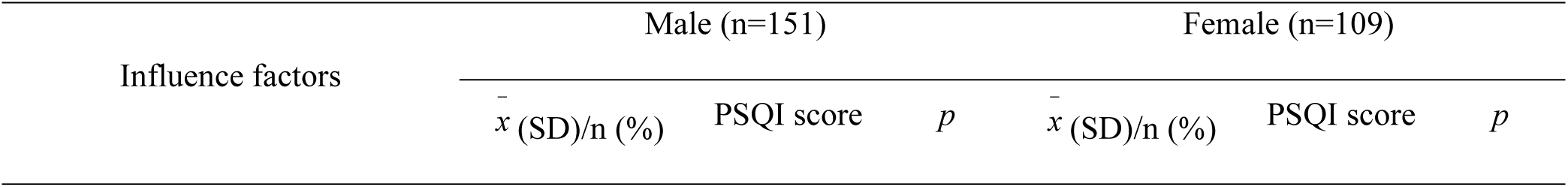
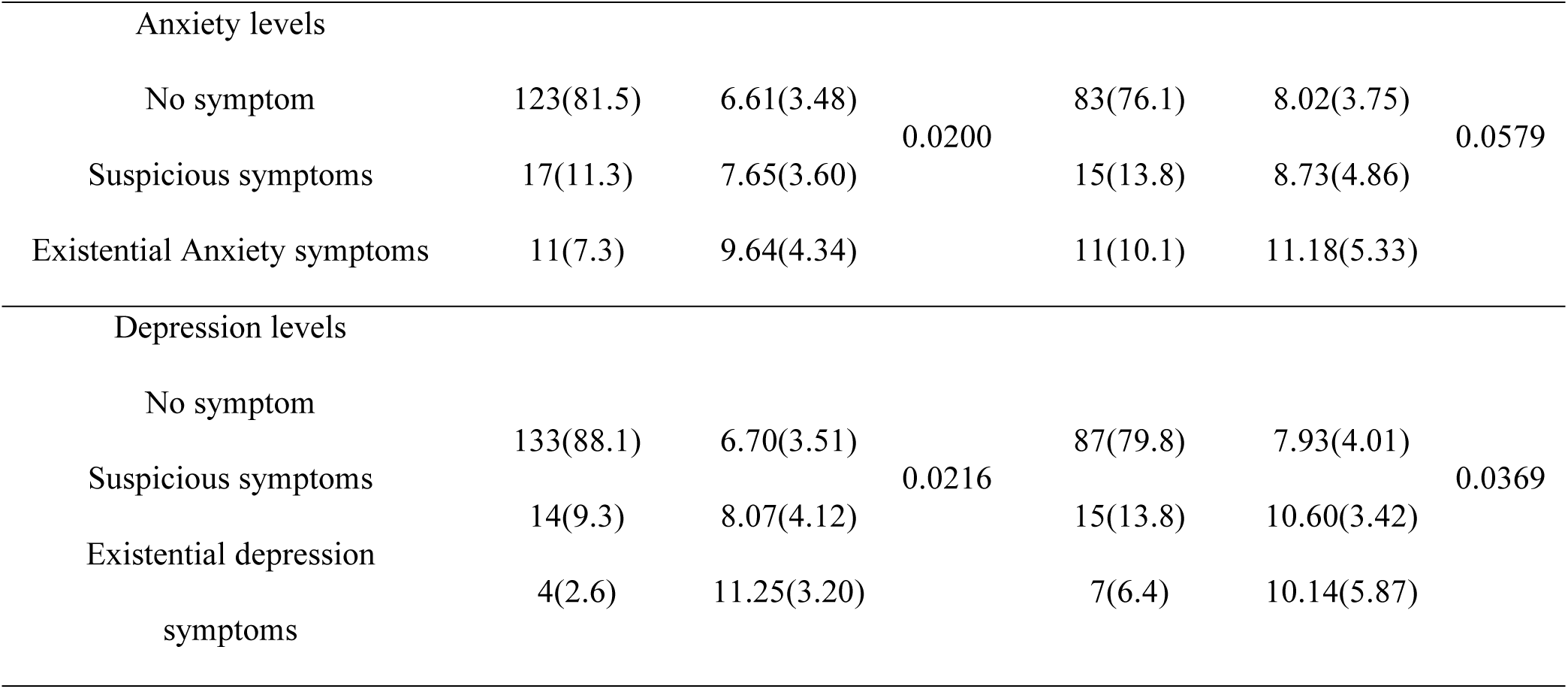
Univariate analysis of emotions and sleep quality.

#### 3.2.3 PSQ is associated with biochemical indicators

To investigate the impact of blood biochemical tests on sleep quality across genders, we retrieved and analyzed the test results from the previous three months (Table 3). For male patients, sleep quality may be linked to ferritin and hemoglobin levels, which are blood biochemical indicators. In contrast, among female patients, sleep quality may be influenced by urea, phosphorus, potassium, and sodium levels, as well as dialysis adequacy in terms of blood biochemical indicators.

**Table 3.** Univariate analysis of biochemical indicators and sleep quality.

| Influence factors | Male (n=151) |  |  | Female (n=109) |  |  |
| --- | --- | --- | --- | --- | --- | --- |
| | $\bar{x}$ (SD) | Estimate | *p* | $\bar{x}$ (SD) | Estimate | *p* |
| Blood uric acid (mmol/L) | 437.98(105.19) | -0.0007 | 0.7994 | 419.55(88.40) | 0.0003 | 0.9481 |
| Urea (mmol/L) | 22.28(6.45) | -0.019 | 0.6741 | 19.77(6.90) | 0.140 | 0.0152 |
| Creatine (umol/L) | 1157.72(288.82) | 0.0001 | 0.8864 | 948.59(245.23) | 0.0003 | 0.8399 |
| eGFR (ml/min/1.73m2) | 4.44(1.75) | -0.029 | 0.8661 | 4.43(1.58) | -0.157 | 0.5388 |
| Total protein (g/L) | 65.78(6.27) | -0.015 | 0.7485 | 67.05(7.24) | 0.007 | 0.9039 |
| Albumin (g/L) | 36.79(4.33) | -0.035 | 0.6065 | 36.27(5.29) | 0.125 | 0.0989 |
| Prealbumin (g/L) | 307.86(71.56) | -0.005 | 0.2022 | 297.52(80.65) | 0.004 | 0.4513 |
| Phosphorus (mmol/L) | 1.97(0.55) | 0.029 | 0.9578 | 1.80(0.53) | 1.593 | 0.0340 |
| Serum potassium (mmol/L) | 4.38(0.73) | -0.206 | 0.6140 | 4.14(0.83) | 0.983 | 0.0408 |
| Sodium (mmol/L) | 139.65(2.63) | -0.139 | 0.2200 | 139.06(3.01) | -0.282 | 0.0337 |
| Chlorine (mmol/L) | 98.70(3.77) | -0.087 | 0.2708 | 97.73(3.63) | -0.124 | 0.2615 |
| Calcium (mmol/L) | 2.20(0.21) | 2.426 | 0.0865 | 2.28(0.21) | 0.892 | 0.6496 |
| β2-microglobulin | 33.16(27.53) | -0.004 | 0.7273 | 36.66(33.93) | -0.009 | 0.4393 |
| Parathyroid hormone (pg/ml) | 49.05(48.12) | 0.005 | 0.3897 | 56.11(71.31) | -0.010 | 0.0861 |
| Dialysis adequacy (Kt/v) | 1.42(0.28) | 0.434 | 0.6833 | 1.77(0.33) | -2.119 | 0.0191 |
| Ferritin (ng/mL) | 193.90(170.05) | 0.005 | 0.0086 | 245.12(223.66) | 0.003 | 0.1625 |
| Hemoglobin (g/L) | 115.40(15.85) | -0.041 | 0.0279 | 110.58(16.42) | 0.021 | 0.3908 |

### 3.3 Multivariate analysis of sleep quality in patients of different sexes

Following stepwise regression analysis, the multivariate linear regression model for the male group incorporated variables such as pruritus (itchy skin), anxiety score, and ferritin. The analysis revealed that in male patients, there was a positive relationship between the presence of itchy skin, anxiety symptoms, and ferritin levels in the blood and the likelihood of developing PSQ after controlling for other variables (Table 4). This model exhibited overall significance, as indicated by the F test, with an F value of 6.546 (*p* < 0.0001).

**Table 4.** Multivariate analysis of sleep quality in male dialysis patients.

| Influence factors | β | Standard error | t value | Confidence interval (95% CI) | p |
| --- | --- | --- | --- | --- | --- |
| Skin itching |  |  |  |  |  |
| No |  |  |  |  |  |
| Yes | 1.850 | 0.567 | 3.265 | (0.730, 2.970) | 0.00136 ** |
| Anxiety levels |  |  |  |  |  |
| No symptom |  |  |  |  |  |
| Suspicious symptoms | 1.111 | 0.956 | 1.162 |  | 0.24714 |
| Existential Anxiety symptoms | 5.045 | 1.734 | 2.910 | (-0.778, 3.000)<br>(1.619, 8.471) | 0.00418 ** |
| Ferritin (ng/mL) | 0.004 | 0.002 | 2.493 | (0.001, 0.007) | 0.01378 * |

Note: \*, $p \leq 0.05$ . \*\*, $p \leq 0.01$ .

Similarly, following stepwise regression screening, the multivariate regression model for the female group included variables such as dialysis modality, systolic blood pressure, depression score, and sodium content. The results indicated that female patients presented an increased risk of sleep disturbances associated with increased reliance on hemodialysis, the presence of depressive symptoms, and elevated systolic blood pressure, even after accounting for other variables (Table 5). The test equation model for this group also displayed overall significance, with an F value of 6.794 (*p* < 0.0001). These findings indicate that reliance on hemodialysis, depression, and uncontrolled hypertension are significant independent predictors of sleep disturbances in female patients even after accounting for other confounding variables in the multivariate model.

**Table 5.** Multivariate analysis of sleep quality in female dialysis patients.

| Influence factors | $\beta$ | Standard error | t value | Confidence interval (95% CI) | *p* |
| --- | --- | --- | --- | --- | --- |
| Dialysis approach |  |  |  |  |  |
| Peritoneal dialysis |  |  |  |  |  |
| Hemodialysis | 2.344 | 0.754 | 3.108 | (0.848, 3.840) | 0.0024 ** |
| Systolic blood pressure (mmHg) | 0.060 | 0.019 | 3.089 | (0.021, 0.098) | 0.0026 ** |
| Depression levels |  |  |  |  |  |
| No symptom |  |  |  |  |  |
| Suspicious symptoms | 0.658 | 1.047 | 0.629 | (-1.418, 2.734) | 0.5310 |
| Existential depression symptoms | 3.700 | 1.198 | 3.088 | (1.324, 6.076) | 0.0026 ** |
| Sodium (mmol/L) | -0.205 | 0.122 | -1.688 | (-0.447, 0.036) | 0.0945 |

Note: \*\*, $p \leq 0.01$ .

### 3.4 The effect of gender on sleep quality in patients on dialysis

After seven risk factors, including age, marital status, pruritus, RLS, systolic blood pressure, depression score, and ferritin, were adjusted for, a pronounced sex difference in the impact on sleep quality outcomes was observed (Table 6). Compared with their male counterparts, female patients manifested heightened vulnerability to PSQ. The female sex appears to be an important nonmodifiable risk factor predisposing individuals to PSQ in this population.

**Table 6.** The impact of gender on sleep quality in dialysis patients.

| Regression model | $\beta$ | Standard error | t value | Confidence interval (95% CI) | $p$ |
| --- | --- | --- | --- | --- | --- |
| Univariate analysis | 1.493 | 0.485 | 3.076 | (0.537, 2.449) | 0.0023 ** |
| Multivariate analysis | 1.184 | 0.447 | 2.651 | (0.304, 2.064) | 0.0085 ** |

Note: \*\*, $p \leq 0.01$ .

## 4 Discussion

Improving sleep quality in patients on dialysis is a pivotal focus of nephrology nursing. While sex differences exist in sleep quality, contemporary strategies for addressing CKD-related PSQ often fail to account for these differences. This study identified noteworthy sex disparities in the risk factors contributing to PSQ among patients on dialysis. For male patients, itchy skin, anxiety, and elevated ferritin levels were significantly associated with PSQ, even after adjusting for other factors. Conversely, hemodialysis use, depression, and increased systolic blood pressure were the primary factors for female patients. These insights pave the way for gender-specific therapeutic interventions and personalized care pathways to optimize sleep quality.

Previous studies have demonstrated the associations of pruritus, depressed mood, and RLS with PSQ in CKD patients [18–20]. Our findings are congruent with this but further emphasize sex- specific variations among patients on dialysis. Specifically, we found that males were more responsive to itching sensations, whereas females displayed greater susceptibility to RLS. Furthermore, anxiety was more prevalent in males, whereas depression was more prevalent in females. Although PSQ is the ultimate outcome, the triggers vary, necessitating distinct treatment approaches. Essentially, many of these symptoms are linked to abnormal neurological functioning.

Extensive research has shown that long-term CKD can lead to neurological complications due to factors such as malnutrition, anemia, toxin accumulation, blood ionic imbalances, and medication side effects [21, 22]. Therefore, in the short term, addressing itching, treating RLS, and providing psychological counseling are potential approaches to alleviate CKD-related PSQ. However, long- term solutions require comprehensive research into the neurological underpinnings of these dysfunctions.

This study also highlighted discrepancies in blood biochemical markers between male and female patients. Specifically, PSQ in female patients was associated with urea, phosphorus, potassium, sodium, and dialysis adequacy. In contrast, for male patients, PSQ was associated with ferritin and hemoglobin levels. Notably, even after multivariate analysis was conducted, ferritin emerged as a significant risk factor for PSQ in male patients, with absolute ferritin levels also being lower in males than in females. Thus, the role of ferritin levels in PSQ has attracted our attention.

Several clinical studies have reported similar results: patients with PSQ generally have low ferritin levels [23, 24]. Ferritin is essential for iron storage and transport in the body, facilitating oxygen delivery within red blood cells and neurotransmitter synthesis. Insufficient levels of ferritin can result in iron deficiency anemia, leading to symptoms such as fatigue, shortness of breath, and heart palpitations, all of which can negatively impact sleep quality. Furthermore, low ferritin levels disrupt neurotransmitter synthesis and function, thereby interfering with the normal regulation of sleep processes. Conversely, iron supplementation has been shown to significantly improve RLS and PSQ [25]. Consequently, we propose that maintaining optimal ferritin levels is imperative for enhancing sleep quality among patients on dialysis. This research direction holds significant importance for our future investigations.

In recent years, the recognition of CKD-related PSQ has increased, leading to the emergence of various nonpharmacologic interventions aimed at improving sleep quality universally. For example, interventions such as increased dietary fiber intake, appropriate melatonin use, and moderate exercise have demonstrated efficacy in enhancing sleep quality, alleviating depressed mood, and enhancing overall quality of life [26–28]. However, few studies have focused specifically on the underlying causes of sex differences in CKD-related PSQ. The findings from this study suggest that nutritional interventions may offer a straightforward and efficacious approach to treatment.

This study does present certain limitations. First, it was confined to a single center, potentially constraining the findings. The exploration of influencing factors has focused primarily on demographic data, dialysis-related factors, and biochemical markers but has not extensively accounted for variables such as psychological status, comorbidities, and lifestyle. For a holistic understanding, future studies should aim to incorporate these additional variables. Second, the sample size was limited, underscoring the need for a larger sample size to bolster statistical significance and generalizability. Third, it is worth mentioning that the prevalence of PSQ in patients on dialysis varies significantly across regions. This discrepancy can be attributed to variations in research tools, ethnicities, and regional cultural differences. Therefore, further research is imperative to understand PSQ and its influencing factors in patients on dialysis across diverse regions. Fourth, employing high-throughput sequencing can be beneficial in comparisons of blood sample differences between sexes. This study aimed to examine the blood components of patients via proteomics and metabolomics. This approach can help identify key factors that significantly impact sleep quality. Fifth, we did not analyze the effects of CKD-related complications on sleep quality. This is partly due to the limited reach of diabetic patients in this study and because CKD patients typically experience numerous complications, leading to a multitude of confounding factors that are challenging to elucidate. Sixth, the type of dialysis affects sleep quality, with hemodialysis being more effective than PD in improving sleep quality. However, due to limitations in the sample size, this study has not conducted further subgroup analyses. Finally, we are currently conducting a study based on sleep polysomnography, which will provide more accurate data for this study. The project is still ongoing due to the volume of work. Such insights are crucial for developing targeted therapies tailored to the specific needs of patients on dialysis.

## 5 Conclusions

In conclusion, there were significant sex differences in the risk factors for CKD-related PSQ, and in general, female patients were more likely to have PSQ. We advocate for the prompt initiation of tailored treatments for CKD patients. For example, male patients might benefit from interventions targeting itchy skin, anxiety management, and iron supplementation. Conversely, addressing depressive symptoms, blood pressure control, and mineral supplementation should be prioritized for female patients. This study has significant implications for the development of nephrology nursing in the future. Future research directions should focus on developing personalized care modalities and treatments.

## Data Availability

The raw data used to support the findings of this study are available from the corresponding author upon request.

## 6 Ethical statement

The research protocols were scrutinized and received approval from the Ethics Committee of Peking University Shenzhen Hospital (Approval No. 2023--066). Before initiation, all participants were verbally informed about the study’s objectives and significance, and their informed consent was duly procured. All patients signed a pan-informed consent form.

## 8 Author Contributions

Clinical data collection: YLiao, YP, and YLiu. Data Analysis: YD, JD, HC, and CL. Research design: YLiao. Writing: YD and GY. Supervision: ZX and YLiu.

## 10 Acknowledgments

We are grateful to all our colleagues in the clinical departments and laboratories.

## Author contributions

Clinical data collection: YLiao, YPeng and YLiu. Data Analysis: YD. Research design: YLiao. Writing: YD and GY. Supervision: GY.

## Supplementary materials

### 13 Supplementary methods

#### 13.1 Patients additional information

Additional information: (a) Patients undergoing peritoneal dialysis received continuous ambulatory peritoneal dialysis throughout the day. (b) Personal information and lifestyle habits of the patients were obtained from medical records or self-reported by the patients themselves, following informed consent. (c) Missing data were either excluded from the statistical analysis or included after employing repeated sampling techniques. (d) The diagnostic criteria for chronic kidney disease (CKD) were based on the 2022 Kidney Disease Improving Global Outcome (KDIGO) guidelines. (e) The classification of the primary disease was determined using outpatient reports. If there were any changes in the diagnostic criteria during treatment, the corresponding data were excluded from the analysis. Following the completion of all the aforementioned steps, the formal study commenced.

#### 13.2 Dialysis

The operating procedures followed the 2021 Standard Operating Procedures. Hemodialysis patients follow a prescribed regimen of 2‒4 dialysis sessions per week to complete treatment. Each treatment session lasted for 4 hours, with a blood flow ranging from 200–300 mL/min and a dialysate flow of 500–600 mL/min. Treatment is administered using either an autologous arteriovenous endovascular fistula or a venous catheter for vascular access. The replacement fluid contained the following concentrations: chloride (118 mmol/L), magnesium (0.797 mmol/L), calcium (1.6 mmol/L), sodium (113 mmol/L), and potassium (adjusted over time). Patients receiving CRRT were not included in this study.

The frequency of peritoneal dialysis ranges from 2 to 5 times a day, with each session involving 2000 ml. The duration of each dialysis session is determined on the basis of the individual patient’s condition, typically lasting 4--6 hours during the day and not exceeding 12 hours at night. For patients experiencing negative ultrafiltration with edema during peritoneal dialysis, the peritoneal fluid is not retained in the abdomen overnight. The peritoneal dialysis fluid utilized was sourced from Baxter®.

#### 13.3 family background

The family background of patients, including employment status, economic status, marital status, and education level, was recorded. In China, CKD is strongly associated with these factors. Hygienic conditions are linked to the living environment and household income, increasing the likelihood of developing infection-related CKD. Additionally, marital status affects mood and postoperative care.

Moreover, patients with low adherence typically have a lower level of education, and they often experience dialysis-related complications due to non-compliance with care. Therefore, these demographic factors were incorporated into this study.

## 14 Supplementary results

### 14.1 Basic personal circumstances

Furthermore, the *p* values for age and marital status are lower for females, and the *p* value for educational attainment is lower for males; however, these *p* values still do not fall below 0.05.

Therefore, it cannot be concluded that these three factors are correlated with PSQ. This could be due to an insufficient sample size and potential survey method bias.

### 14.2 Lifestyle habits

We assessed the correlation between sleep quality and various lifestyle habits, including the consumption of alcohol, tea, and coffee, and exercise frequency among patients of different sexes (Table S2). However, the data did not reveal any significant associations between these habits and sleep quality. The lack of associations between these variables and sleep quality warrants further investigation in larger patient samples.

**Table S1.** Univariate analysis of basic personal information and sleep quality.

| Influence factors | Male (n=151) |  |  | Female (n=109) |  |  |
| --- | --- | --- | --- | --- | --- | --- |
| | $\bar{x}$ ( SD) /n (%) | PSQI score | *p* | $\bar{x}$ ( SD) /n (%) | PSQI score | *p* |
| Age (years) | 52.21(13.03) | 6.95(3.63) | 0.2361 | 50.50(13.29) | 8.44(4.16) | 0.0914 |
| Education |  |  |  |  |  |  |
| ≤ Middle school | 43(28.5) | 7.16(3.29) | 0.1914 | 60(55.0) | 8.47(4.51) | 0.9739 |
| High school | 52(34.4) | 7.50(4.03) |  | 25(22.9) | 8.28(3.49) |  |
| ≥ College degree | 56(37.1) | 6.27(6.27) |  | 24(22.2) | 8.54(4.04) |  |
| Marital status |  |  |  |  |  |  |
| Unmarried | 11(7.3) | 6.45(3.93) | 0.5528 | 5(4.6) | 5.20(1.79) | 0.1158 |
| Married | 133(88.1) | 6.92(3.65) |  | 96(88.1) | 8.72(4.20) |  |
| Divorce | 6(3.9) | 8.83(2.93) |  | 4(3.7) | 9.00(4.32) |  |
| Widow | 1(0.7) | 5.00(-) |  | 4(3.7) | 5.25(2.87) |  |
| Working condition |  |  |  |  |  |  |
| Full-time | 41(27.2) | 6.20(2.91) | 0.4651 | 12(11.0) | 8.42(3.45) | 0.2486 |
| Half position retire | 26(17.2) | 6.77(3.60) |  | 18(16.5) | 8.22(3.73) |  |
| Retire | 50(33.1) | 7.22(3.88) |  | 33(30.3) | 9.73(4.64) |  |
| Retirement | 9(6.0) | 6.89(4.14) |  | 11(10.1) | 8.27(3.07) |  |
| Unemployment | 25(16.6) | 7.84(4.05) |  | 35(32.1) | 7.40(4.29) |  |
| Monthly income |  |  |  |  |  |  |
| 1000-3000 RMB | 38(25.2) | 7.16(4.39) | 0.8203 | 61(56.0) | 8.08(4.49) | 0.4944 |
| 3000-5000 RMB | 53(35.1) | 7.17(3.58) |  | 25(22.9) | 9.00(3.64) |  |
| 5000-1 RMB | 33(21.9) | 6.76(3.04) |  | 21(19.3) | 9.10(3.86) |  |
| Over 10000 RMB | 27(17.9) | 6.44(3.36) |  | 2(1.8) | 5.50(0.71) |  |

**Table S2.** Univariate analysis of lifestyle habits and sleep quality.

| Influence factors | Male (n=151) |  |  | Female (n=109) |  |  |
| --- | --- | --- | --- | --- | --- | --- |
| | $\bar{x}$ ( SD) /n (%) | PSQI score | *p* | $\bar{x}$ ( SD) /n (%) | PSQI score | *p* |
| Drink alcohol |  |  |  |  |  |  |
| Yes | 3 | 3.33(3.06) | 0.0800 | 0 | - | - |
| No | 148 | 7.02(3.61) |  | 109 | 8.44(4.16) |  |
| Drink tea |  |  |  |  |  |  |
| Yes | 78(51.7) | 6.95(3.66) | 0.9953 | 24(22.0) | 8.92(4.43) | 0.5278 |
| No | 73(48.3) | 6.95(3.63) |  | 85(78.0) | 8.31(4.10) |  |
| Drink coffee |  |  |  |  |  |  |
| Yes | 20(13.2) | 6.90(3.31) | 0.9507 | 3(2.8) | 9.00(3.46) | 0.8145 |
| No | 131(86.8) | 6.95(3.69) |  | 106(97.2) | 8.42(4.19) |  |
| Sports |  |  |  |  |  |  |
| Yes | 122(80.8) | 6.88 (3.61) | 0.6289 | 80(73.4) | 8.40(4.05) | 0.8673 |
| No | 29(19.2) | 7.24 (3.79) |  | 29(26.6) | 8.55(4.53) |  |

## References

1. Topal Kılıncarslan, G., A. Özcan Algül, and N. Gördeles Beşer, Sleep quality, coping, and related depression: A cross-sectional study of Turkish nurses. Int Nurs Rev, 2024.

2. Hodgson, N.A., et al., Timed Activity to Minimize Sleep Disturbance in People With Cognitive Impairment. Innov Aging, 2024. 8(1): p. igad132.

3. Tan, L.H., et al., Insomnia and Poor Sleep in CKD: A Systematic Review and Meta-analysis. Kidney Med, 2022. 4(5): p. 100458.

4. Peng, Y., et al., Risk factors affecting the sleep quality of patients on dialysis: A single-center cross-sectional study. Medicine (Baltimore), 2024. 103(13): p. e37577.

5. Hosseini, M., et al., Relationship of sleep duration and sleep quality with health-related quality of life in patients on hemodialysis in Neyshabur. Sleep Med X, 2023. 5: p. 100064.

6. Yazıcı, R. and İ. Güney, Prevalence and related factors of poor sleep quality in patients with pre-dialysis chronic kidney disease. Int J Artif Organs, 2022. 45(11): p. 905–910.

7. Kennedy, C., et al., The impact of change of renal replacement therapy modality on sleep quality in patients with end-stage renal disease: a systematic review and meta-analysis. J Nephrol, 2018. 31(1): p. 61–70.

8. Yang, G., et al., COVID-19 increases extracorporeal coagulation during hemodialysis associated with upregulation of vWF/FBLN5 signaling in patients with severe/critical symptoms. BMC Infectious Diseases, 2024. 24(1): p. 427.

9. Liao, Y., et al., Involuntary Falls in Patients with Chronic Kidney Diseases on Nephrology Wards: Research Advances and Future Perspectives. Nursing: Research and Reviews, 2024. 14: p. 1–12.

10. Krishnan, V. and N.A. Collop, Gender differences in sleep disorders. Curr Opin Pulm Med, 2006. 12(6): p. 383–9.

11. Amiri, S. and S. Behnezhad, Sleep Disturbances and Physical Impairment: A Systematic Review and Meta-Analysis. Physical & Occupational Therapy In Geriatrics, 2021. 39(3): p. 258–281.

12. Turnbull, C.D., et al., Relationships between MRI fat distributions and sleep apnea and obesity hypoventilation syndrome in very obese patients. Sleep Breath, 2018. 22(3): p. 673–681.

13. Gaultney, J.F., P. Zendels, and A. Ruggiero, Gender differences affecting the relationship between sleep attitudes, sleep behaviors and sleep outcomes. Cogent Psychology, 2021. 8(1): p. -.

14. von Elm, E., et al., The Strengthening the Reporting of Observational Studies in Epidemiology (STROBE) statement: guidelines for reporting observational studies. J Clin Epidemiol, 2008. 61(4): p. 344–9.

15. Ware, J., Jr., M. Kosinski, and S.D. Keller, A 12-Item Short-Form Health Survey: construction of scales and preliminary tests of reliability and validity. Med Care, 1996. 34(3): p. 220–33.

16. Buysse, D.J., et al., The Pittsburgh Sleep Quality Index: a new instrument for psychiatric practice and research. Psychiatry Res, 1989. 28(2): p. 193–213.

17. Razavi, D., et al., Screening for adjustment disorders and major depressive disorders in cancer in-patients. Br J Psychiatry, 1990. 156: p. 79–83.

18. Nevols, J., L. Watkins, and R. Lewis, A phase IV, randomised, double-blind, controlled, parallel group trial to evaluate the effectiveness and safety of Balneum Plus versus emollient in the treatment of chronic kidney disease-associated pruritus in haemodialysis patients. Clin Kidney J, 2023. 16(8): p. 1307–1315.

19. Burton, J.O., et al., Current Practices in CKD-Associated Pruritus: International Nephrologist Survey. Kidney Int Rep, 2023. 8(7): p. 1455–1459.

20. Chen, J., et al., Network of depressive symptoms before and after a diagnosis of chronic kidney disease. Health Psychol, 2023.

21. Hilderman, M. and A. Bruchfeld, The cholinergic anti-inflammatory pathway in chronic kidney disease-review and vagus nerve stimulation clinical pilot study. Nephrol Dial Transplant, 2020. 35(11): p. 1840–1852.

22. Arnold, R., et al., Neurological complications in chronic kidney disease. JRSM Cardiovasc Dis, 2016. 5: p. 2048004016677687.

23. Benbir Senel, G., et al., Restless sleep disorder in children with epileptic and non-epileptic nocturnal attacks. J Sleep Res, 2023.

24. Thorarinsdottir, E.H., et al., Serum ferritin and obstructive sleep apnea-epidemiological study. Sleep Breath, 2018. 22(3): p. 663–672.

25. Leung, W., et al., Iron deficiency and sleep - A scoping review. Sleep Med Rev, 2020. 51: p. 101274.

26. Zhang, S., et al., Association of dietary fiber with subjective sleep quality in hemodialysis patients: a cross-sectional study in China. Ann Med, 2023. 55(1): p. 558–571.

27. Yousef, E.A., et al., Study of Exogenous Melatonin as a Treatment Modality for Sleep and Psychiatric Disorders in Hemodialysis Patients. Saudi J Kidney Dis Transpl, 2022. 33(1): p. 1–15.

28. Luo, Y., et al., Effect of a video-based exercise intervention on depression and sleep conditions of peritoneal dialysis patients. Clin Nephrol, 2023. 99(2): p. 58–68.

